# CHA2DS2-VASc score on admission to predict mortality in COVID-19 patients: A meta-analysis

**DOI:** 10.1101/2021.06.07.21258402

**Authors:** Prattay Guha Sarkar, Amit Kumar, Debdutta Bandyopadhyay

## Abstract

**Background:** CHA2DS2-VASc score is used in non-valvular AF patients to predict thromboembolic risk. Recent studies have tried to evaluate CHA2DS2-VASc score on admission in COVID-19 patients to predict mortality.

**Methods:** We conducted a literature search on 14 April 2021 to retrieve all published studies, pre-prints and grey literature related to the predictive power of CHA2DS2-VASc score in COVID-19 patients of admission and mortality. Screening of studies and data extraction was done by two authors independently. We used the Quality in Prognosis Studies (QUIPS) tool for the methodological quality assessment of the included studies.

**Results:** Five studies involving 5,941 patients reported the predictive value of CHA2DS2-VASc score for mortality in COVID-19 patients. The pooled sensitivity (SEN), specificity (SPE) and area under curve were 0.72 (95% CI 0.63-0.79), 0.74 (95% CI 0.67-0.81) and 0.80 (95% CI 0.76-0.83).

**Conclusions:** CHA2DS2-VASc score at admission has good predictive value for mortality in patients with COVID-19 infection and can help clinicians identify potentially severe cases early. Early initiation of effective management in these cases may help in reducing overall mortality due to COVID-19.

**Trial registry:** We prospectively registered this meta-analysis on PROSPERO database (Reg number: CRD42021248398).

## Introduction

The ongoing COVID-19 pandemic is one of the most severe crises faced by healthcare systems worldwide. Respiratory failure is one of the most common reasons for admissions into critical care units. However, SARS-CoV-2 infection has proven to be a complex disease with multi-organ involvement in many cases. Hypoxia and generalized endothelial dysfunction due to viral invasion leading to thromboembolic complications has been postulated as a common mechanism of multi-organ failure^1^. Mortality has been significantly higher in elderly patients with pre-existing cardiovascular risk factors^2^. Poorer outcomes in these patients may be caused by the higher incidence of endothelial dysfunction and thromboembolism leading to multi-organ failure^3^.

The CHA2DS2-VASc score is a clinical score used patients of non-valvular AF to predict the risk of thromboembolism^4^. In addition, this score has been also used to predict mortality in other cardiovascular condition without AF. This score has a prognostic significance independent of AF^5^ as it is a cluster of common cardiovascular risk factors seen in clinical practice.

CHA2DS2-VASc score can be readily calculated bedside using its components C(Congestive Heart Failure), H(Hypertension), A2(Age>75 years; 2 points), D(Diabetes Mellitus), S2(Stroke, TIA or thromboembolism; 2 points), V(vascular disease), A(Age 65-74 years) and Sc(Female sex)^4^. We performed this meta-analysis to evaluate the predictive values of CHA2DS2-VASc score in COVID-19 patients for mortality so as to identify potentially high risk cases early.

## Methods

The Preferred Reporting Items for Systematic Reviews and Meta-Analyses (PRISMA statement) guidelines were followed to perform this meta-analysis^6^. We prospectively registered this meta-analysis on PROSPERO database (Registration number: CRD42021248398).

### Selection of studies

We reviewed PubMed, Google scholar, Scirius, Medline, Liliacs, Cochrane, CINAHIL, Plos and SIGLE databases through April 14, 2021. The search terms were as follows: “2019 Coronavirus disease” or “Novel Coronavirus” or “SARS-CoV-2-19” or “2019-nCoV” or “COVID-19” and “CHA2DS2-VASc score”. Language restrictions were avoided. Additional citations were identified by screening the reference list of the included studies.

Twoauthors (P.G.S and D.B) independently identified the studies to be included into the final analysis. Any disagreement was resolved through discussion. In case of persistent disagreement, a third reviewer (A.K) was consulted for arbitration. Studies were selected if the following criteria were met (1) The predictive value of CHA2DS2-VASc score on mortality in patients of COVID-19 was evaluated; (2) Sufficient information was available to calculate a 2×2 table for true positive (TP), false positive (FP), true negative (TN) and false negative (FN). Exclusion criteria were (1) Unable to extract 2×2 table; (2) Case reports, reviews, comment, letter, animal studies.

### Data extraction and quality assessment

Relevant information from individual studies was extracted by two independent authors (P.G.S and D.B). Extracted information included area under curve (AUC), cut-off value, TP, TN, FP, FN, sensitivity (SEN) and specificity (SPE). Third author (A.K) checked all the extracted information. We used the Quality in Prognosis Studies (QUIPS) tool to assess risk of bias (ROB) in 6 domains: participation, attrition, prognostic factor measurement, outcome measurement, confounding factors and statistical analysis & reporting.^7^

### Statistical analysis

We used random effects model to compute the pooled sensitivity, pooled specificity with 95% CI considering the significant heterogeneity among the studies. Summary area under the curve was computed to determine the discriminating power of CHA2DS2-VASc score for mortality. Diagnostic odds ratio was computed to provide the accuracy of CHA2DS2-VASc score for the predicting mortality. Heterogeneity more than 50% was considered as statistically significant heterogeneity. Mete-regression analysis was done to determine the source of heterogeneity and subgroup effects. All the statistical analyses was completed using software STATA version 13.

## Results

### Selection and characteristic of studies

Figure 1 shows the study selection process. We reviewed PubMed, Google scholar, Scirius, Medline, Liliacs, Cochrane, CINAHIL, Plos and SIGLE databases through April 14, 2021 and identified 26 studies. An additional 11 records were identified through other sources. 9 records found in duplicates were removed. Abstracts of the remaining 28 studies were scrutinized thoroughly. 19 studies were excluded as they did not report predictive value of CHA2DS2-VASc in COVID-19 patients. Full text articles of 9 studies were evaluated. 3 studies had reported CHA2DS score and 1 study had reported m-CHA2DS2-VASc score, hence were excluded. Finally, five studies reporting the predictive value of CHA2DS2-VASc score on mortality in COVID-19 patients, were included in this meta-analysis.

**Fig 1:**
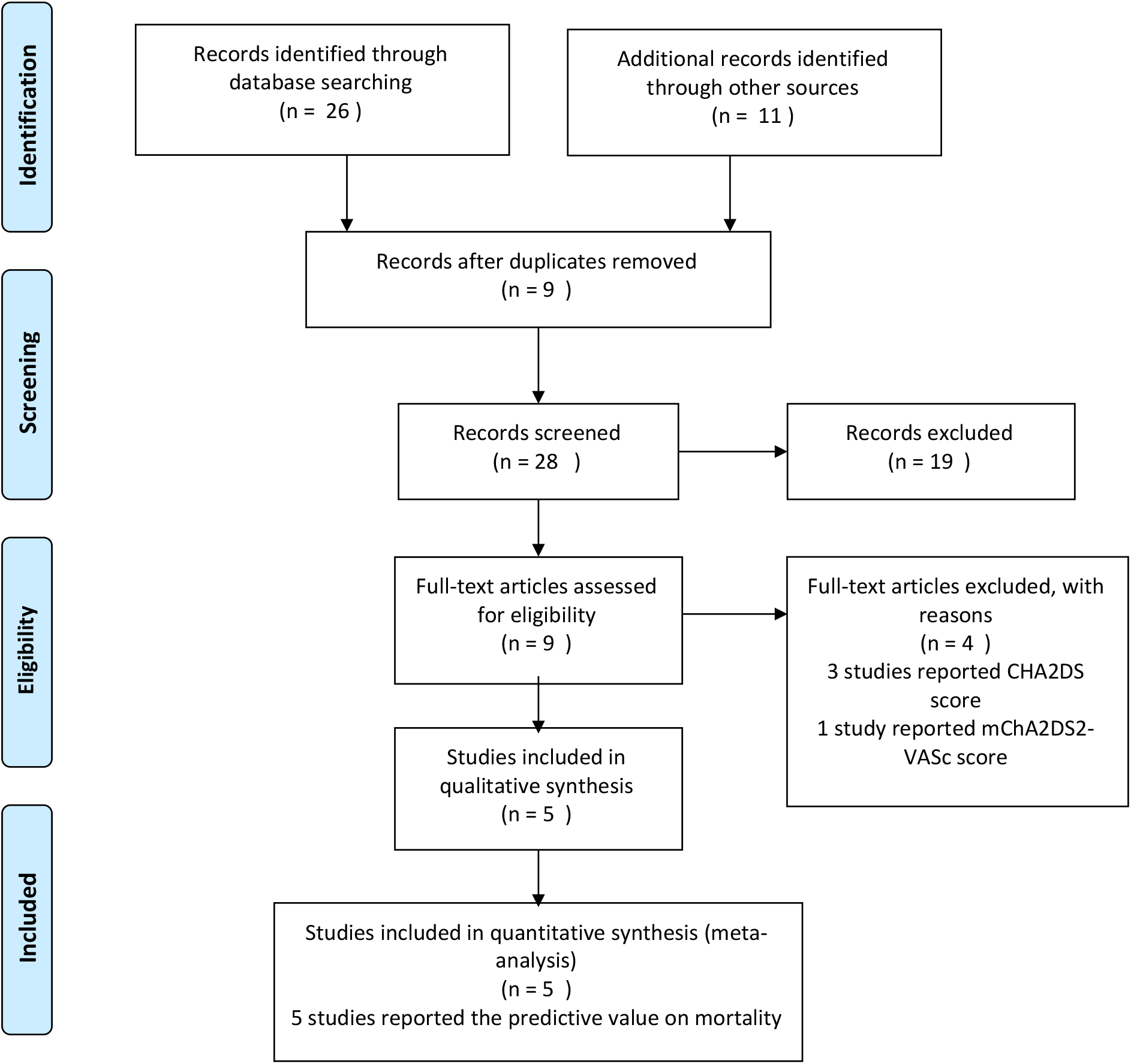
Flow diagram for the identification of eligible studies.

The characteristics of each study and the predictive value of CHA2DS2-VASc score for mortality in COVID-19 patients are presented in Table 1. Three studies have been conducted in Turkey, one in Spain and one in Italy. All the studies were retrospective in nature. The number of participants varied from 318 to 3042. All the studies have reported SEN, SPE and AUC for CHA2DS2-VASc score for predicting mortality in COVID-19 patients.

**Table 1:** Characteristics of the included studies and diagnostic test performance of CHA2DS2-VASc score in predicting mortality in COVID-19

|  |  |  |  |  |  |
| --- | --- | --- | --- | --- | --- |
| RUOCCO G | GUNDUZ R | CICEK V | CARO-CODON J | CAETINKAL G | FIRST AUTHOR |
| 2020 | 2021 | 2020 | 2021 | 2020 | YEAR |
| ITALY | TURKEY | TURKEY | SPAIN | TURKEY | STUDY LOCATION |
| MORTALITY | MORTALITY | MORTALITY | MORTALITY | MORTALITY | GROUP |
| 62.2 | 56.9 | 78.1 | 63.3 | 57.6 | MALE % |
| 65 +- 17.0 | NA | 75 +- 13.3 | 80.5 +-11.0 | 64 +-10.97 | AGE |
| 136 (15.7) | 69 (33) | 13 (40.6) | 196 (31.3) | 187 (26.9) | DM N(%) |
| 420 (48.6) | 148 (70.8) | 25 (78.1) | 437 (69.8) | 349 (50.2) | HTN (%) |
| 53 (6.1) | 15 (7.2) | NA | 74 (11.8) | 42 (6) | CHF N(%) |
| 65 | 81.71 | 69 | 75 | 60 | SENSITIVITY |
| 73 | 83.88 | 78 | 75 | 60 | SPECIFICITY |
| 0.69 | 0.8907 | 0.75 | 0.794 | 0.64 | AUC |
| 2.5 | 2.5 | 2.5 | NA | NA | CUT OFF |
| 697 | 918 | 286 | 2416 | 618 | SURVIVOR |
| 167 | 82 | 32 | 626 | 76 | NON SURVIVOR |

### Study quality and publication bias

The methodological quality of the included studies is presented in Fig.2 and Fig.3. Risk of Bias assessment was done by QUIPS tool. Risk of Bias domains evaluated include participation, attrition, prognostic factor measurement, outcome measurement, confounding factors and statistical analysis & reporting. Studies by Caetinkal^8^ et al and Cicek^9^ et al were found to have high risk of confounding factors as it was not clearly mentioned how the assessment was done. Study by Gunduz^10^ et al was found to have high risk of attrition as control population follow up was not mentioned. Studies by Caro-codon^11^ et al and Ruocco^12^ et al scored low to moderate in various Risk of Bias domains. We could not do publication bias analysis due to insufficient number of studies to conduct such analysis.

**Fig 2:**
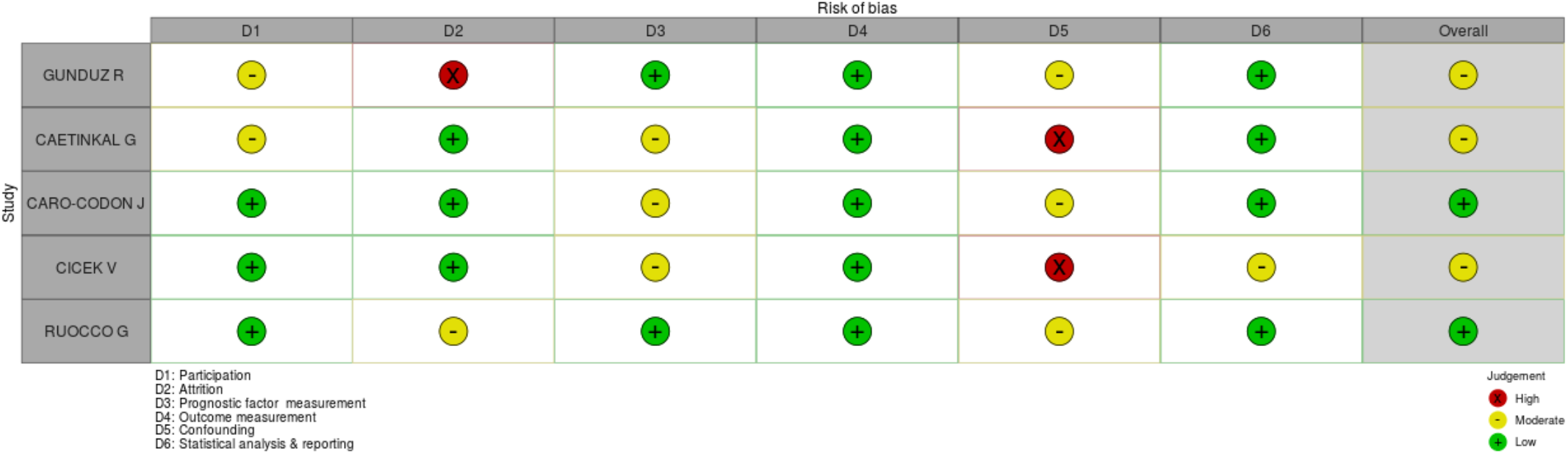
Risk of Bias assessment of individual studies by QUIPS tool.

**Fig 3:**
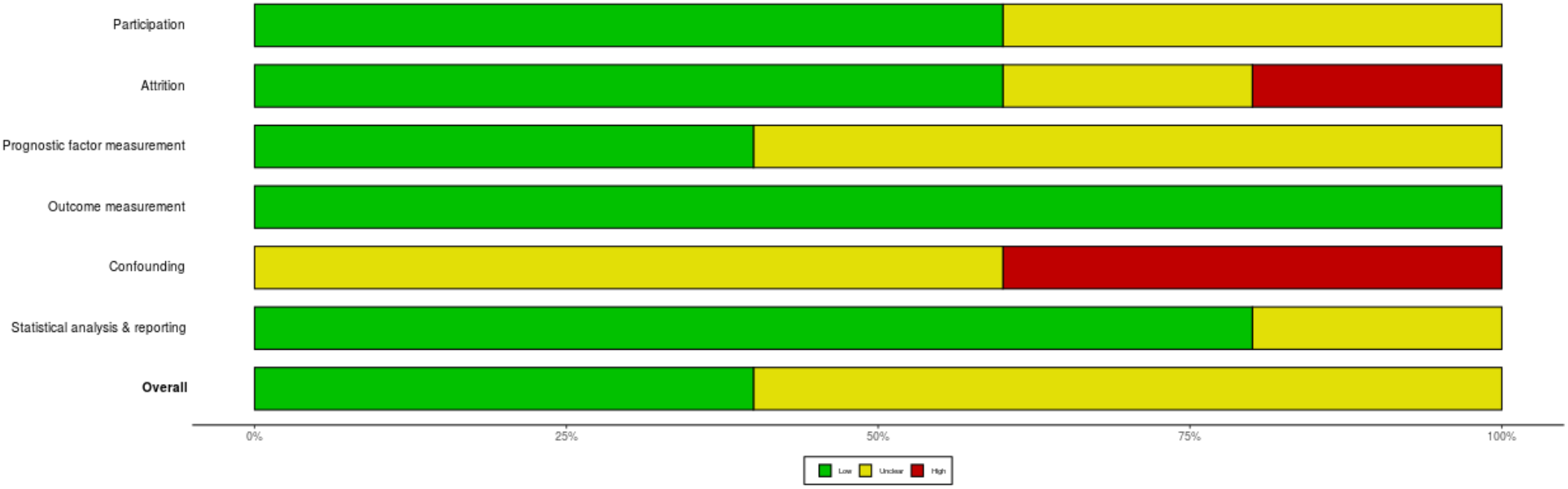
Overall Risk of Bias assessment by QUIPS tool.

### Predictive value of CHA2DS2-VASc score on mortality

Five studies involving 5,941 patients reported the predictive value of CHA2DS2-VASc score for mortality in COVID-19 patients. The pooled sensitivity (SEN) and specificity (SPE) were 0.72 (95% CI 0.63-0.79) and 0.74 (95% CI 0.67-0.81), respectively (Fig.4). The positive likelihood ratio was 2.8 (95% CI 2.0-4.0) and the negative likelihood ratio was 0.37 (95% CI 0.26-0.54). The diagnostic odds ratio (DOR) was 8 (95% CI 4-15). The pooled AUC of CHA2DS2-VASc score for discriminating mortality was 0.80 (95% CI 0.76-0.83), indicating that it has high predicting accuracy (Figure 5). Fagan normogram (Fig.6) shows that if the pre-test probability was set to 20%, the post-test probability of CHA2DS2-VASc score for predicting mortality was 41% when the score was 2.5. When the CHA2DS-VASc score was below cut-off value, post-test probability was only 8%.

**Fig.4:**
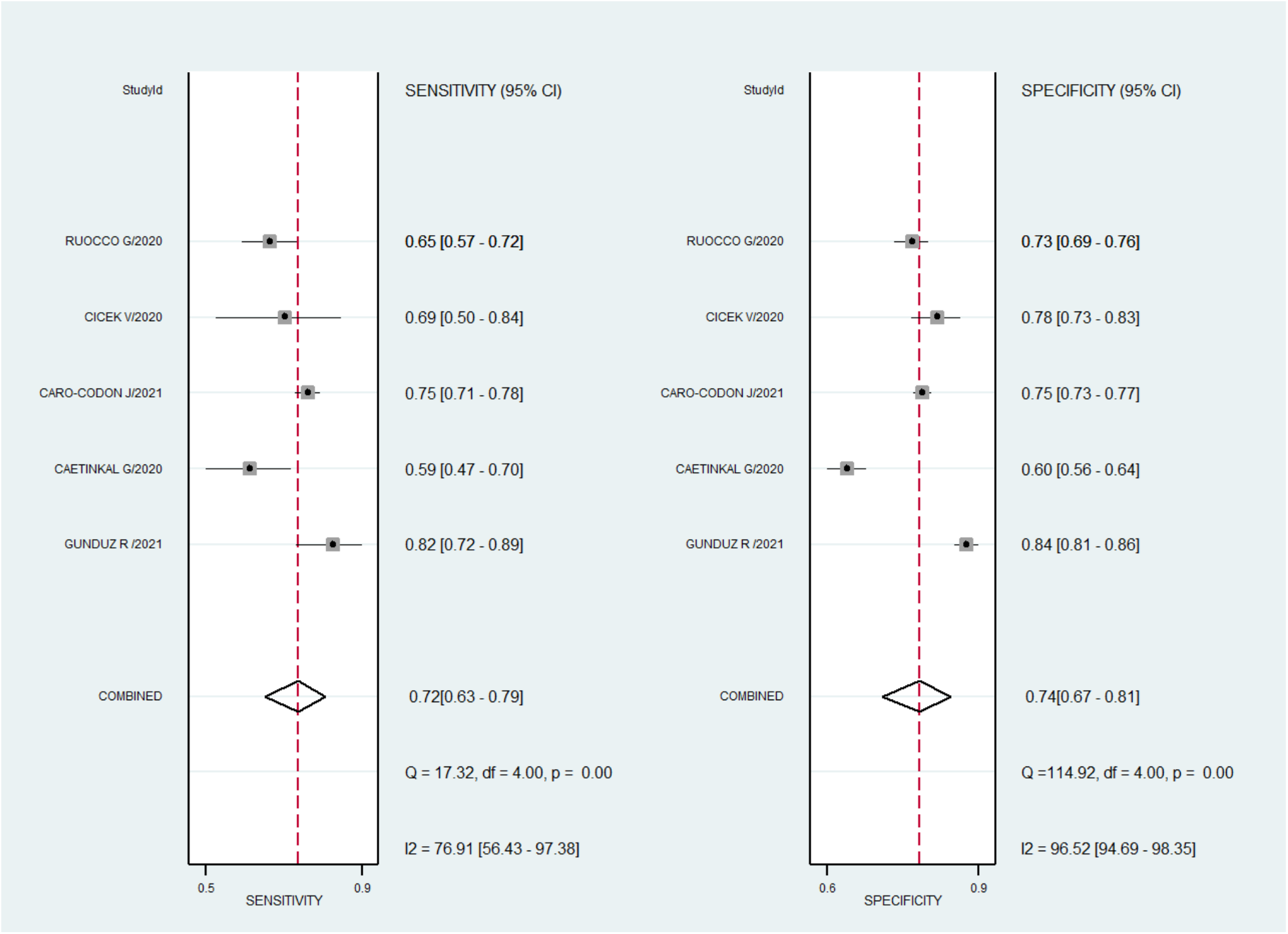
Forest plot of the sensitivity and specificity of CHA2DS2-VASc score to predict mortality in COVID -19 patients. The pooled sensitivity (SEN) and specificity (SPE) were 0.72 (95% CI 0.63-0.79) and 0.74 (95% CI 0.67-0.81).

**Fig.5:**
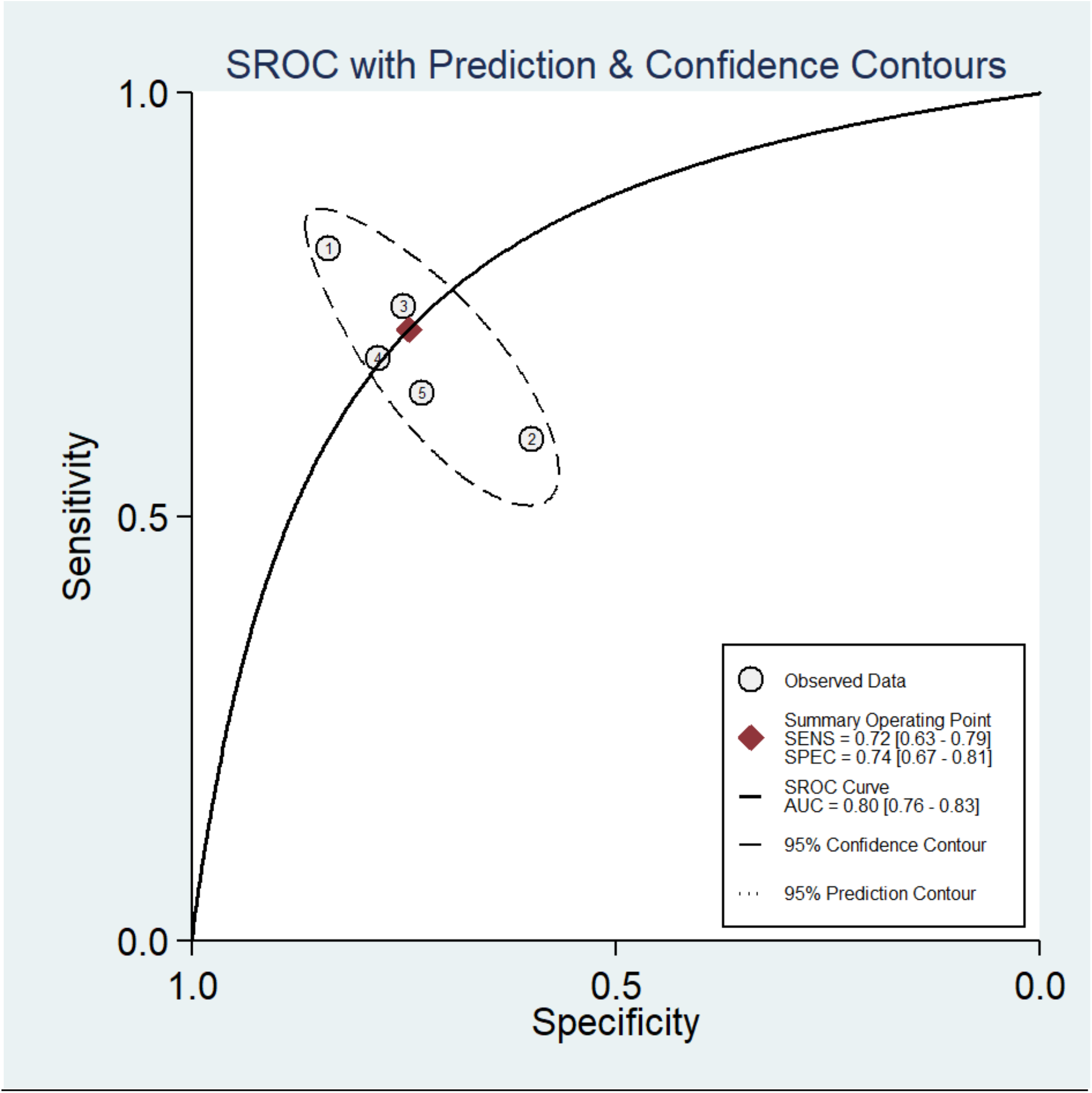
Summary receiver operating characteristic graph of the included studies. The AUC of CHA2DS2-VASc score for predicting mortality was 0.80 (95% CI 0.76-0.83).

**Fig.6:**
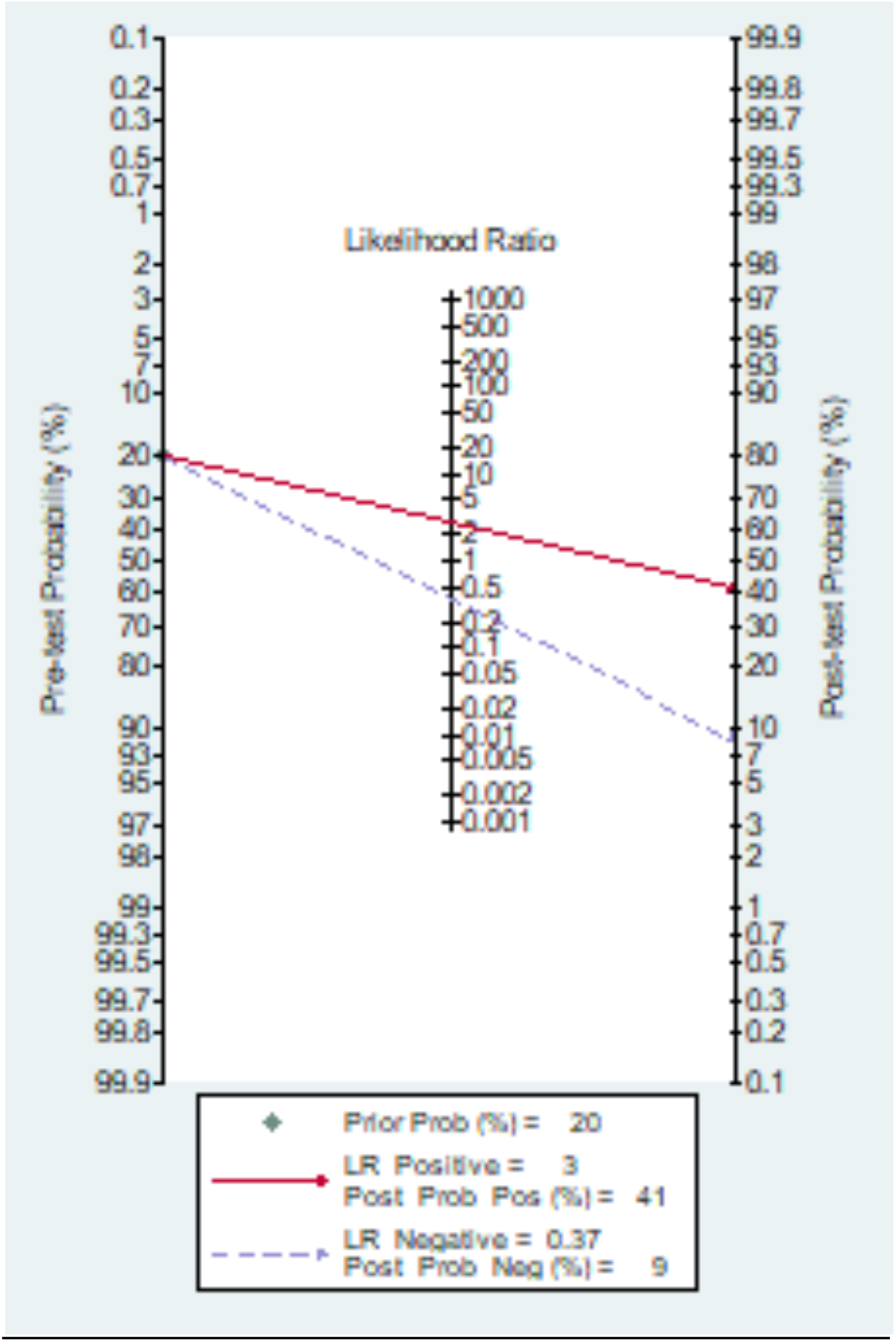
Fagan normogram of CHA2DS2-VASc score to predict mortality in COVID-19 patients.

**Fig.7:**
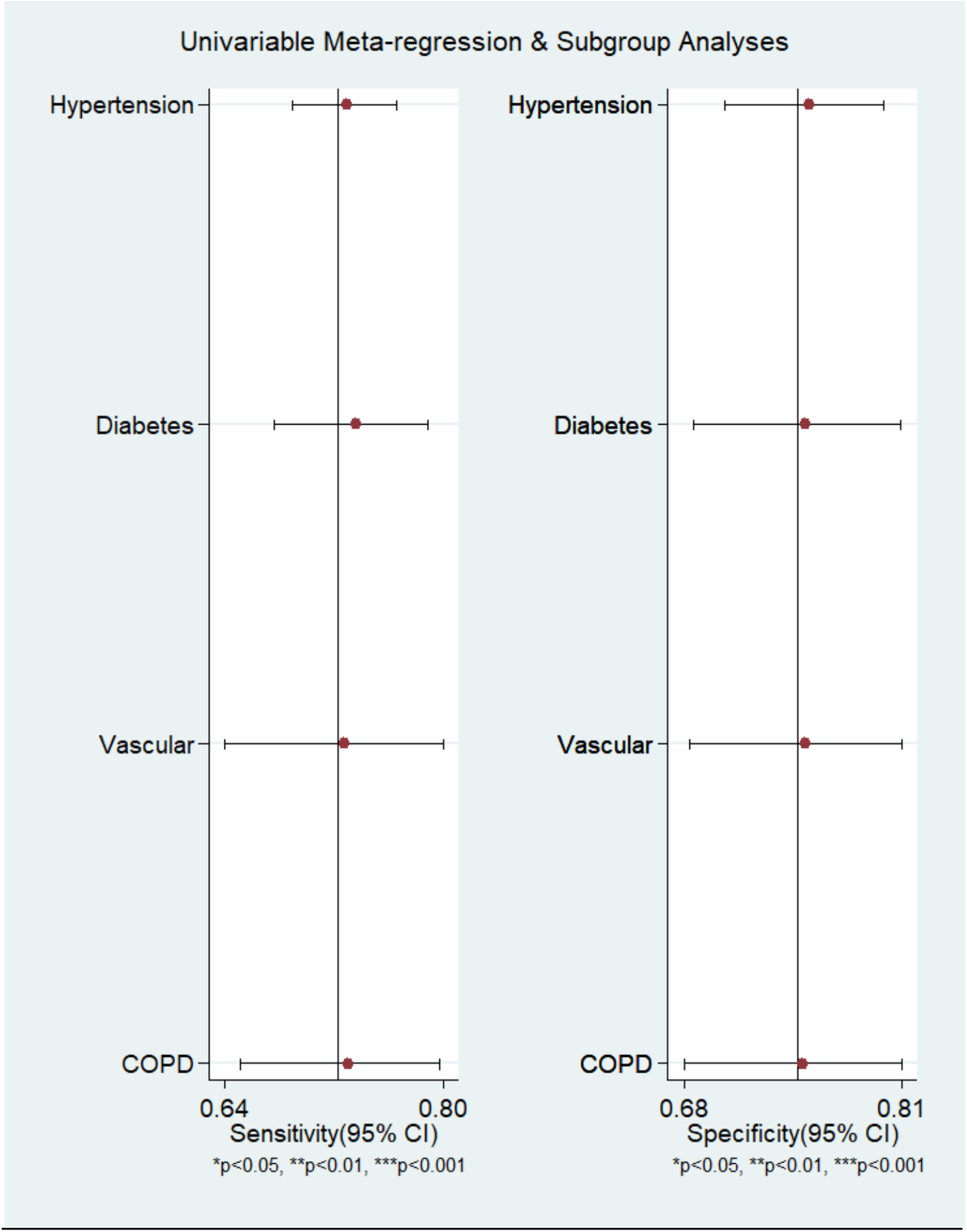
Meta-regression analysis no statistically significant covariate effects of hypertension, diabetes, vascular risk factor and COPD on the pooled sensitivity and pooled specificity.

### Subgroup analyses

We did the meta-regression analysis to determine the covering factors on summary measure performance. Only four variables (hypertension, diabetes, vascular risk factor and COPD) had sufficient data to conduct meta-regression analysis. We observed that no statistically significant covariate effects of hypertension, diabetes, vascular risk factor and COPD on the pooled sensitivity and pooled specificity. Although, small number of studies limiting power of regression to detect significant effects indicating that more studies are required to detect such effects if any.

## Discussion

Covid-19 pandemic poses a significant stress on healthcare system of the entire world. In the present scenario, it is very important to improve triage and risk stratification and to provide clinicians with easily accessible tools to easily predict adverse clinical outcomes. Scoring systems like APACHE II (Acute physiology and chronic health evaluation), COVID-GRAM etc are being studied to predict disease progression and outcome^13^. However, both these scores are difficult to be applied in resource limited settings as they are heavily dependent on laboratory examinations. APACHE II calculations need pH and serum electrolytes, COVID-GRAM calculation needs lactate dehydrogenase, both of which may not be available in a resource limited setting^14^. Therefore, simpler clinical tools to predict progression and mortality in COVID-19 patients in early stages is urgently needed.

In clinical practice with COVID-19 patients, we have observed an increased incidence of mortality due to thromboembolism predicted by CHA2DS2-VASc score. There is no systematic review and meta-analysis to evaluate the predictive value of CHA2DS2-VASc score for mortality in COVID-19 patients. Studies have reported various threshold for the same. Clinicians are therefore unsure of the sensitivity and specificity of the score to predict mortality in COVID-19 patients. In our final analyses, five studies reporting CHA2DS2-VASc score to predict mortality in COVID-19 patients have been included. The pooled sensitivity (SEN) and specificity (SPE) were 0.72 (95% CI 0.63-0.79) and 0.74 (95% CI 0.67-0.81), respectively. The AUC of CHA2DS2-VASc score for predicting mortality was 0.80 (95% CI 0.76-0.83), indicating that it has high prognostic value.

COVID-19 has been described as a hyper-inflammatory state leading to diffuse pulmonary intravascular coagulopathy^15^. Excess mortality in these patients is often related to progressive respiratory failure and thromboembolism. The CHA2DS2-VASc score is widely used to stratify thromboembolic risk in AF patients. In addition, they also predict all-cause mortality among a wide spectrum of cardiovascular disorders without AF. CHA2DS2-VASc score has been used to predict mortality in COVID19 patients too. Most of the variables of the CHA2DS2-VASc score such as elderly age, diabetes, and hypertension are also confirmed prognostic risk factors in patients hospitalized with COVID-19. Because CHA2DS2-VASc score clubs common risk factors predicting a pro-thrombotic, we can expect higher mortality in COVID-19 cases with higher CHA2DS2-VASc score. Therefore, this is the first reported meta-analyses to report that a simple bedside clinical score, CHA2DS2-VASc score, can be used to predict mortality in COVID-19 patients with high prognostic value.

This meta-analysis has the following shortcomings. First, all the studies are retrospective in nature, hence prone to confounding factors. Second, prognosis of the disease may be affected by other factors not directly included in the score. Third, as only five studies were available for meta-analysis, publication bias and sub-group analyses could not be performed. Additional high-quality studies may be required to further shed light on the prognostic significance of this score in COVID-19 patients.

## Conclusion

Evaluating CHA2DS2-VASc score can help clinicians identify severe cases early. Initiation aggressive and timely management in these high risk patients can help in reducing mortality due to COVID-19.

## Data Availability

The authors confirm that the data supporting the findings of this study are available within the article. The studies included for synthesis of this meta-analysis have been mentioned in references.

## Notes

The authors have no conflicts of interest.

### Competing Interest Statement

The authors have declared no competing interest.

### Clinical Trial

PROSPERO CRD42021248398

### Clinical Protocols

https://www.crd.york.ac.uk/prospero/display_record.php?RecordID=248398

### Funding Statement

NONE

